# Effects of exercise, metformin, and their combination on glucose metabolism in individuals with impaired glycemic control: A systematic review and network meta-analysis

**DOI:** 10.1101/2024.01.22.24301604

**Authors:** Tong Zhao, Qize Yang, Joshua F. Feuerbacher, Bizhu Yu, Christian Brinkmann, Sulin Cheng, Wilhelm Bloch, Moritz Schumann

## Abstract

**Objective:** To compare the efficacy of exercise, metformin, and their combination on glucose metabolism in individuals with prediabetes and type 2 diabetes mellitus (T2DM), and rank these treatments by exercise modality and metformin dosage.

**Design:** Systematic review and network meta-analysis.

**Data sources:** Embase, Web of Science, PubMed/MEDLINE, and SPORTDiscus were searched until February 2023.

**Eligibility criteria for selecting studies:** Randomized controlled trials (RCTs) of exercise, metformin, or their combined treatments in individuals with prediabetes or T2DM were included.

**Analyses:** We estimated pooled mean difference (MD) with 95% confidence intervals (CI) for five glucose metabolism variables (i.e., hemoglobin A1c [HbA1c], 2-h glucose during oral glucose tolerance test [OGTT], fasting glucose, fasting insulin, and homeostasis model assessment of insulin resistance [HOMA-IR]) via a random effect model. Subgroup analyses were conducted for prediabetes and T2DM populations.

**Results:** We identified 15,872 eligible articles and finally included 375 articles with 378 RCTs, comprising 30,884 participants. When all individuals were pooled, metformin had greater effects than exercise in improving HbA1c (MD −0.65 95% CI [−0.77 to −0.53] vs −0.38 [−0.45 to −0.32] %), 2-h glucose during OGTT (−1.16 [−1.67 to −0.65] vs −0.76 [−1.15 to −0.37]), fasting glucose (−0.99 [−1.14 to −0.83] vs −0.57 [−0.65 to −0.48] mmol/L), and fasting insulin (−2.28 [−3.03 to −1.53] vs −1.47 [−1.85 to −1.09] μU/mL), but not in HOMA-IR (−0.36 [−0.77 to 0.04] vs −0.73 [−0.90 to −0.57]). A similar pattern was observed for the subgroup of T2DM patients; however, in prediabetes, exercise showed better efficacy than metformin in HbA1c (−0.17 [−0.23 to −0.11] vs −0.09 [−0.20 to 0.01] %) and 2-h glucose (−0.68 [−1.01 to −0.35] vs −0.04 [−0.51 to 0.43] mmol/L). Exercise + metformin showed a greater efficacy than exercise alone in improving HbA1c (−0.79 [−1.27 to −0.30] %) and fasting glucose (−0.76 [−1.25 to −0.26] mmol/L) when all individuals were pooled. Considering exercise modalities, aerobic interval exercise was most efficient in improving fasting glucose (−0.82 [−1.06 to −0.59] mmol/L), HbA1c (−0.61 [−0.77 to −0.44] %), fasting insulin (−2.22 [−3.34 to −1.10] μU/mL), and HOMA-IR (−0.95 [−1.39 to −0.51]). The confidence in evidence was mostly low or very low.

**Conclusion:** The use of exercise, metformin, and their combination are all effective in improving glucose metabolism in individuals with impaired glycemic control, such as prediabetes or T2DM, but the efficacy varies in the five outcome parameters (i.e., HbA1c, OGTT 2-h glucose, fasting glucose, fasting insulin, and HOMA-IR). The efficacy was modified by exercise modality, metformin dosage, and severity of impaired glycemic control. Future clinical trials may further investigate the specific components of the interactive effects of exercise and metformin, such as the timing of exercise and metformin administration, the drug delivery method as well as the effect of additional exercise variables (e.g., exercise frequency and volume).

PROSPERO registration number: CRD42023400622.

## INTRODUCTION

Diabetes is one of the most common chronic diseases worldwide, which has considerable impacts on well-being of individuals and societies.^1^ In 2019, 463 million people (9.3% of the world population) lived with diabetes and the number is projected to exceed 700 million (10.9%) in 2045.^2^ Furthermore, according to the criteria of the American Diabetes Association (ADA),^3^ one-third of the US population is in a prediabetes state.^4^ Approximately three-quarters of individuals with prediabetes at age 45 years have a likelihood of developing type 2 diabetes mellitus (T2DM) during their lifetime.^5^ Thus, it is essential to give due attention to the impaired glycemic control, even in the early phases.

Metformin and exercise training are both considered as first-line treatments for T2DM,^6,7^ and are recommended in those with prediabetes to delay the development of diabetes.^8^ Since being marketed, metformin has been widely used in clinical practice, because it is effective, safe, and affordable.^9^ Meanwhile, increasing evidence emphasizes the benefits of physical exercise training for glycemic control and overall well-being.^10^ Although the mechanisms of metformin and exercise seem different, both may activate the key regulatory enzyme AMPK and elicit similar effects on glucose metabolism, such as increased glucose uptake and insulin sensitivity.^11^ Thus, the combination of exercise and metformin is hypothesized to have a higher efficacy than each standalone treatment and hence is commonly prescribed in individuals with impaired fasting glucose or glucose tolerance.^12^ However, there are several controversies in terms of exercise and metformin treatments in individuals with impaired glycemic control.

First, several studies opposed the use of metformin in individuals with prediabetes,^13^ where regular physical exercise has been recommended as an alternative treatment with no major side effects and pharmaceutical expenditure.^14^ However, the existing meta-analyses regarding the efficacy of exercise and metformin in prediabetes only concentrated on straightforward binary outcomes (i.e., mortality and incidence of diabetes).^15–17^ Thus, there is a current absence of comprehensive comparisons of exercise and metformin for the entire glucose metabolism profile, which is essential to support clinical practice in the prediabetes population.

Second, despite the well-documented benefits of exercise training on glycemic control, the effects and mechanisms of different exercise modalities are inconsistent.^10^ Previous meta-analyses have compared the effects of different exercise modalities in glycemic control, yet they used different classifications of exercise modalities (e.g. comparing supervised versus unsupervised training or only focusing on aerobic versus resistance exercise but without specifying intensity domains) and exclusively concentrated on individuals diagnosed with T2DM.^18–20^ Thus, it remains elusive as to which is the most efficient exercise modality for the population with varying degrees of impaired glycemic control (i.e., prediabetes and T2DM).

Third, although exercise and metformin are commonly prescribed together for glycemic control, several studies found no additive effects of “exercise + metformin” compared to each treatment alone.^21,22^ Contrary, some studies even showed blunted improvements in glucose metabolism when metformin and exercise were prescribed concomitantly.^23,24^ Furthermore, the recent consensus from the American College of Sports Medicine (ACSM) highlighted the inter-individual variability in the efficacy of the exercise-metformin interaction,^10^ which may be affected by exercise modality.^11^ However, to date a systematic evaluation on the comparative efficacy of regular physical exercise and metformin alone, compared to their combination on glucose metabolism is lacking.

To partly account for these gaps, this study utilizes a systematic review and network meta-analysis to provide a holistic synthesis of the role of exercise, metformin, and their combination for indices related to glucose metabolism (i.e., hemoglobin A1c [HbA1c], 2-h glucose during oral glucose tolerance test [OGTT], fasting glucose, fasting insulin, and homeostasis model assessment of insulin resistance [HOMA-IR]) in individuals with impaired glycemic control. Specifically, the following three research questions were assessed: (1) Is exercise as efficient as metformin in improving glucose metabolism? (2) Which type of exercise is most efficient? (3) Is the combination of exercise and metformin more effective than each treatment alone? In addition, subgroup analyses were performed assessing these research questions also in individuals with prediabetes or diagnosed T2DM.

## METHODS

### Search strategy and selection criteria

This review was registered with PROSPERO (CRD42023400622) and designed in accordance with the guidelines of the preferred reporting items for systematic reviews and meta-analyses (PRISMA) checklist for network meta-analyses (Appendix 1).^25^ Only peer-reviewed original human studies in English since 1990 were included. Four databases were searched: Embase, Web of Science, PubMed/MEDLINE, and SPORTDiscus. We developed a systematic search strategy to identify studies by using the terms *“Exercise” OR “Metformin” AND“Glucose metabolism”* (Appendix 4).

Eligibility criteria were defined according to PICOS (Population, Intervention, Comparator, Outcomes, and Study design). The *population* included adults (age≥18 years) with impaired glycemic control (i.e., prediabetes and T2DM). Prediabetes was defined as individuals whose mean baseline glucose parameters met at least one of the following criteria: fasting glucose 5.6–6.9 mmol/L, 2-h glucose 7.8–11.0 mmol/L, HbA1c 5.7%–6.4%.^3^ T2DM was defined as fulfilling one of the following criteria: fasting glucose≥7.0 mmol/L, 2-h glucose≥11.1 mmol/L, HbA1c≥6.5%, random plasma glucose≥11.1 mmol/L.^3^ There were no restrictions to sex and body mass index (BMI). The *intervention* consisted of (1) any type of structured exercise training, (2) metformin treatment within therapeutic doses (i.e., 500–2550 mg/day),^26^ or (3) their combination. For *comparison*, the reported effects were examined as follows: (1) exercise vs control, (2) metformin vs control, (3) exercise + metformin vs control, (4) exercise + metformin vs exercise, (5) exercise + metformin vs metformin, or (6) exercise vs metformin. Parameters of the glucose metabolism (i.e., HbA1c, 2-h glucose during OGTT, fasting glucose, fasting insulin, and HOMA-IR) were used as *outcomes*. The *study designs* were restricted to randomized controlled trials (RCTs), including cluster-randomized or crossover designs. Studies were excluded if they (1) focused on pregnant women, type 1 diabetes, and patients presenting acute medical conditions (e.g., patients in the hospital emergency department); (2) consisted of acute interventions (e.g., a single bout of exercise/ dose of metformin), and (3) interventions combined with diet, insulin, or other interventions.

Exercise was defined as a subset of planned, structured, and repetitive physical activity for improving or maintaining physical fitness.^27^ We categorized the exercise intervention into six types: (1) aerobic continuous exercise, (2) aerobic interval exercise (i.e. including any exercise intensity, e.g. high-intensity interval exercise [HIIT]), (3) resistance exercise, (4) mind-body exercise (e.g., Tai-Chi, yoga), and (5) combined exercise (i.e., combinations of the exercise types mentioned), and (6) other exercises, similar to a previous review (detailed definitions are listed in Appendix 3).^28^ Metformin treatment was divided into low- and high-doses (i.e., 500–1500 mg/day, and >1500 mg/day, respectively).^29^ Exercise and metformin co-intervention was further categorized as (1) aerobic exercise + metformin, (2) resistance exercise + metformin, and (3) combined exercise + metformin.

The search results were imported into the systematic review tool Rayyan (https://rayyan.ai/). After removing duplicates, the records were screened based on title and abstract. The full texts were then retrieved and assessed for inclusion in the review.

### Data extraction and processing

We extracted information on study characteristics (i.e., first author name, publication year), participant characteristics (i.e., sample size, age, sex, baseline symptoms severity), intervention protocol, outcome measures (i.e., HbA1c [%], OGTT 2-h glucose [mmol/L], fasting glucose [mmol/L], fasting insulin [µU/mL], and HOMA-IR). For the outcomes reported at multiple time points, we recorded the measures as close to 8 weeks as possible for all analyses. If the information at 8 weeks was not available, we gave preference to the timepoint closest to 8 weeks; if equidistant (e.g., 4 weeks and 12 weeks), we took the longer outcome.^30^ For studies reporting both intention-to-treat (ITT) and per-protocol (PP) analysis results, we used the PP analysis results as the majority of the included trials reported PP but not ITT analysis.

For the calculation of pooled effects, we extracted means and standard deviations (SDs) of post-intervention measurement or change-from-baseline outcomes value, as well as the number of participants in each intervention group. If median, standard errors, 95% confidence intervals (CI), interquartile ranges, or ranges were reported, these were transformed into means and SDs using established methods (Appendix 5).^31^ If the data was presented in graphs, the WebPlotDigitizer (Version 4.6.0) was used to extract the underlying numerical data. If the required information was not provided, the corresponding authors were contacted to request the data. Otherwise, the missing SDs were estimated by utilizing the mean SD of the remaining studies.^31^ For studies with multiple similar interventions, the data from the multiple groups were combined by using an established method (Appendix 5).^31^ The data from cluster-randomized and crossover design trials were treated by using methods from the Cochrane Handbook (Appendix 5).^31^ For multiple reports of the same trial, we selected the articles with the most informative (i.e., reporting more outcome parameters) and complete (i.e., including larger participant sample) data.

### Assessment of risk of bias and evaluating confidence in the evidence

We assessed the risk of bias by using version 2 of the Cochrane risk-of-bias (RoB 2) tool, which includes important innovations in the assessment of the risk of bias in randomized trials.^32^ The Confidence in Network Meta-Analysis (CINeMA, https://cinema.ispm.unibe.ch/) was used to assess the confidence of the network meta-analysis.

Two researchers (TZ and QY) independently screened the studies, extracted the data, and assessed the risk of bias of included studies. Any disagreements between the two researchers were resolved by discussion with a third author (MS).

### Statistical analysis

We conducted network meta-analyses in R (Version 4.2.3) package *netmeta*.^33^ The key codes of analyses are provided in Appendix 6. All analyses were performed based on two-level classification of interventions: general classification (Classification 1: exercise, metformin, and exercise + metformin) and detailed classification (Classification 2: different types of exercise, different doses of metformin, and different types of exercise + metformin; see detailed classification above).

We estimated pooled mean differences (MDs) and 95% CIs from outcomes by performing random effects pairwise and network meta-analyses (for direct and indirect comparisons) with an inverse variance model. We used Cochran’s *Q* test to estimate between-study heterogeneity, and qualified the effect of heterogeneity as *I*^2^ (the percentage of total variation across studies that is due to heterogeneity rather than chance; 25%, 50%, and 75% indicated low, medium, and high levels of heterogeneity, respectively).^34^ We presented the MDs from pairwise and network meta-analyses using the thickness of lines in network graphs and forest plots (*forestploter* package), respectively. Furthermore, we used the *P*-score (with 1 indicating the best and 0 indicating the worst) to rank the treatments for each outcome if applicable.

We assessed transitivity in the network by comparing variables (and their distributions) that may act as effect modifiers: publication year of included studies, intervention duration, and participant characteristics (i.e., age of participants, proportion of women, and baseline outcome measures). Inconsistency in the network was measured with local and global approaches: the former used the separate indirect from direct evidence (SIDE) method in *netsplit* function, to assess inconsistency between direct and indirect evidence within each comparison (i.e., loop inconsistency); the latter used *Q* statistics based on a full design-by-treatment interaction random effect model in *decomp.design* functions, to assess both loop inconsistency and inconsistency stemming from study designs (i.e., design inconsistency). The publication bias and small study bias were assessed by funnel plots and Egger’s test.

We also performed subgroup analyses by using different populations (i.e., prediabetes and T2DM) as a covariate, to evaluate the potential heterogeneity of treatment effects in these two populations. Sensitivity analysis was conducted after the exclusion of trials at high risk of bias and trials whose intervention periods are longer than 12 weeks.

### Equity, diversity, and inclusion statement

Our research team included various genders, countries, and career stages from multiple disciplines (i.e., physical training, physiology, endocrinology, and clinical medicine). Our study population included both males and females without regional, educational, and socioeconomic restrictions.

## RESULTS

After removing duplicates, a total of 15,872 potentially eligible records were identified. Of 1,170 records retrieved for full-text screening, 375 studies were finally included in the network based on inclusion criteria (*n*=378 RCTs, with multiple sub-trials within one study, Figure 1). The degrees of agreement between the two independent reviewers during abstract and full-text screening were substantial (kappa score=0.70) and almost perfect (kappa score=0.94), respectively. The detailed study selection process is presented in Appendix 7.

**Figure 1.**
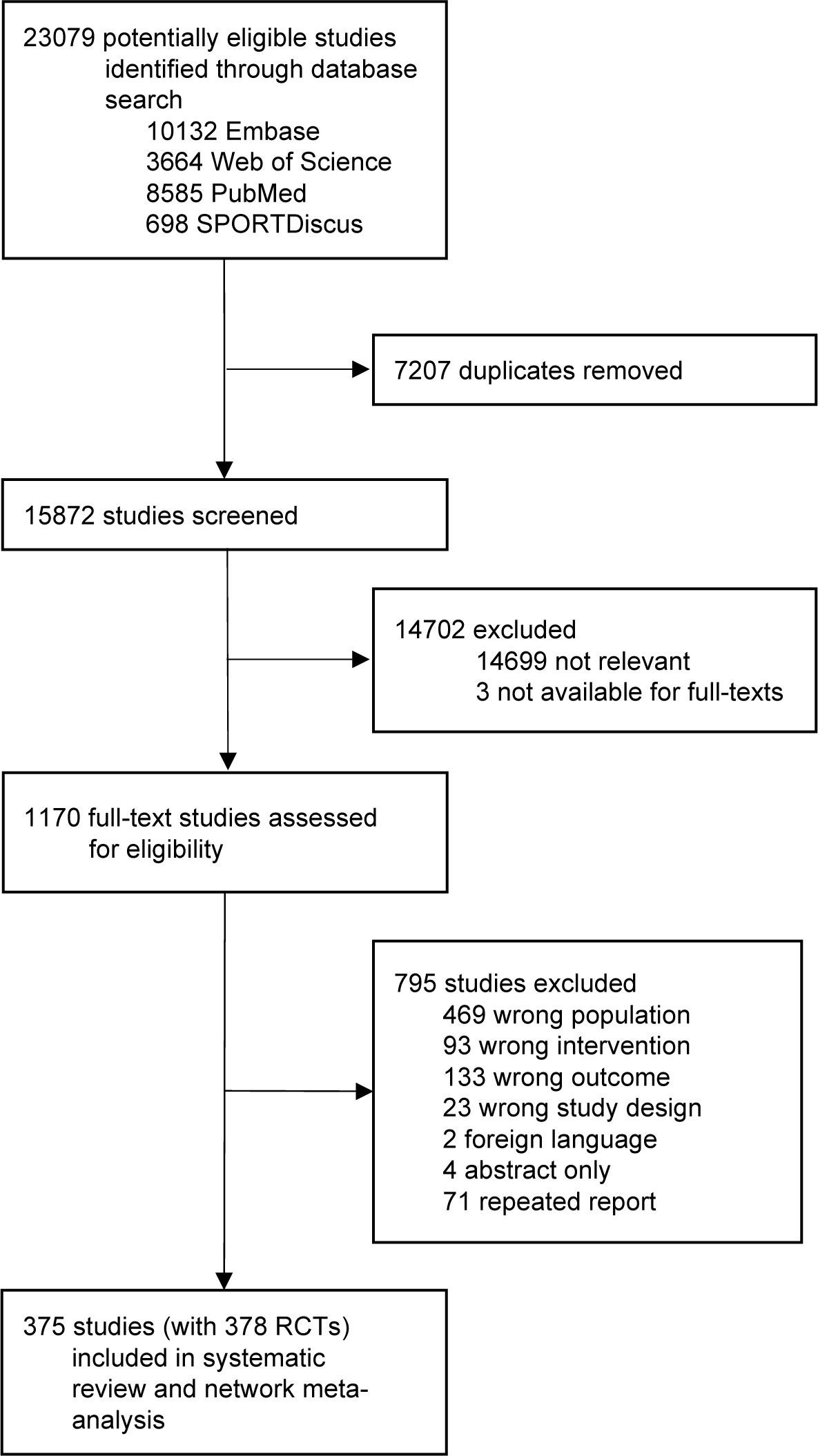
Study identification and screening process. *Note.* Of 378 RCTs of included studies, 297 were exercise interventions, 76 were metformin interventions, and 5 were combined exercise and metformin interventions. RCT=Randomized controlled trial.

### Characteristics of included studies

Overall, 30,884 participants (11,539 prediabetes and 19,345 T2DM) were randomized into the control (*n*=13,469) and one or more of the following treatment groups: aerobic continuous exercise (*n*=3,736), aerobic interval exercise (*n*=1,045), resistance exercise (*n*=1,673), mind-body exercise (*n*=1,011), combined exercise (*n*=2,853), other exercises (*n*=362), low-dose metformin (*n*=3,210), high-dose metformin (*n*=3,430), aerobic exercise + metformin (*n*=60), resistance exercise + metformin (*n*=19), and combined exercise + metformin (*n*=16). The mean age of participants was 56.5 years (SD 9.0), 52.8% were female, and 47.2% were male. The main characteristics of the included studies and participants are summarized in Appendix 8.

Transitivity assessment for all included studies (i.e., prediabetes and T2DM) revealed no significant difference among treatment groups in terms of participant characteristics (i.e., age and baseline outcome measures), except for the proportion of female participants (Appendix 15). In the risk of bias assessment, 13 (3.4%) were rated high risk, 97 (25.7%) with some concerns, and 268 trials (70.9%) at low risk (Appendix 9). Funnel plots and Egger’s test suggested a small-study bias on outcomes of fasting glucose (*p*<0.001) and HbA1c (*p*<0.001). No further small-study effect was shown in other outcome variables (Appendix 16).

## Main analyses

### HbA1c

For the changes in HbA1c of the pooled population, 271 RCTs were included in the network with 215 comparisons for exercise, 59 for metformin, and 3 for exercise + metformin (Figure 2A). The results of the pairwise meta-analysis comparing with the control group revealed that standalone metformin (Figure 2A and Appendix 10; MD −0.66 95% CI [−0.78 to −0.54] %) had larger effects on HbA1c than exercise (−0.38 [−0.45 to −0.31] %), while there was no study directly comparing exercise + metformin and control. The network meta-analysis showed that exercise + metformin (Figure 3 and Appendix 10; −0.79 [−1.27 to −0.30] %) led to larger reductions in HbA1c than metformin (−0.65 [−0.77 to −0.53] %) and exercise alone (−0.38 [−0.45 to −0.32] %). The network meta-analysis, in detailed classification based on exercise modalities and metformin dosages (i.e., Classification 2), found that all types of included treatments decreased HbA1c significantly, with MD ranging from −1.37 (−2.14 to −0.61) to −0.26 (−0.46 to −0.07) % (Figure 3). However, there was a lack of data on combined exercise + metformin. The ranking on HbA1c level indicated resistance exercise + metformin as the best and resistance exercise alone as the worst treatment (Figure 4; Appendix 13).

**Figure 2.**
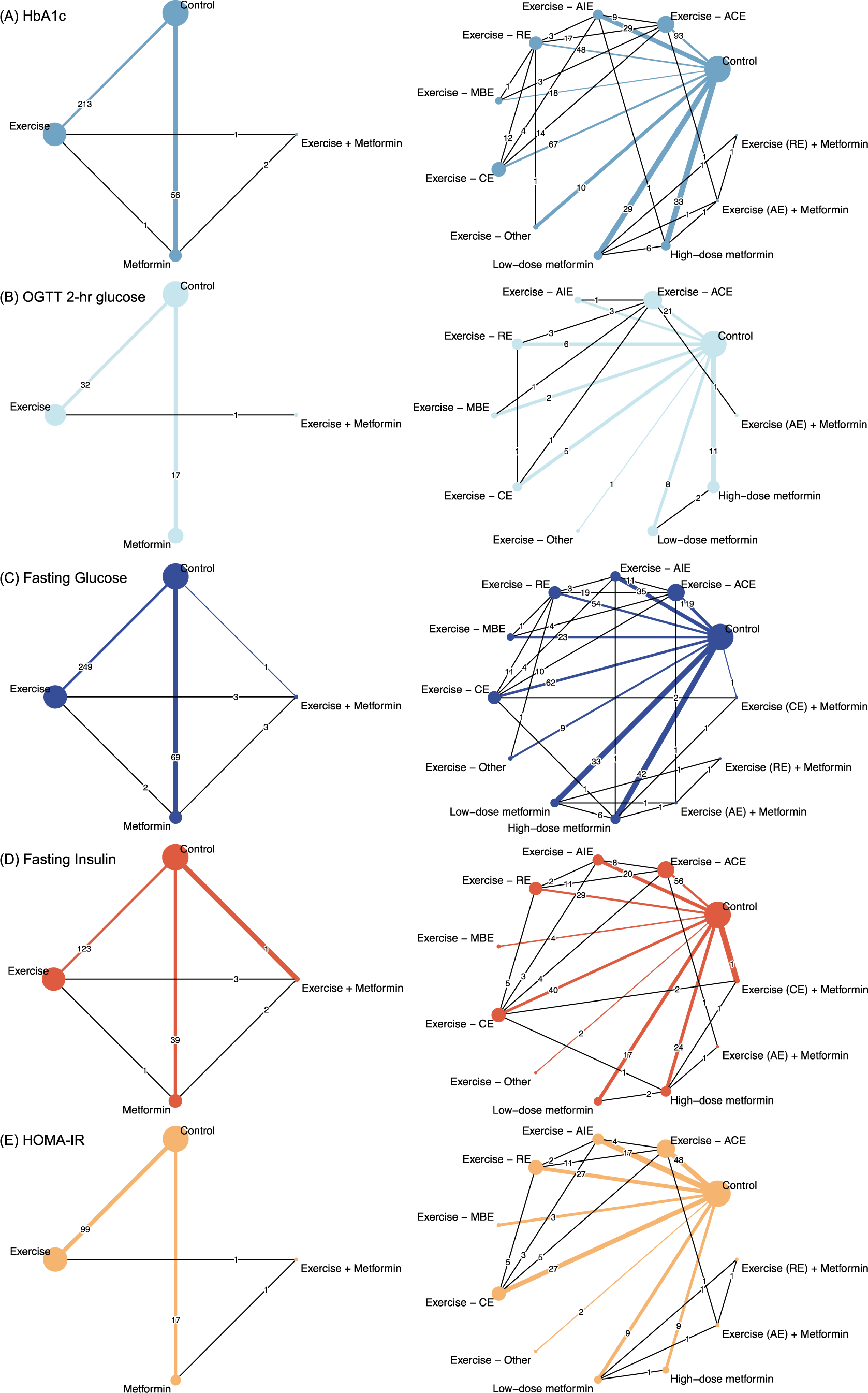
Network of eligible comparisons for glucose metabolic parameters. *Note.* The pooled effects (i.e., mean differences) of pairwise meta-analyses (direct comparisons) are shown as the thickness of the lines in the network graphs (see details in Appendix 10). Left column (classification 1): all exercise, metformin, their combination, and control condition; right column (classification 2): exercises in different types, metformin in different doses, their combination, and control condition. ACE=aerobic continuous exercise; AIE=aerobic interval exercise; CE=combined exercise; HOMA-IR=homeostatic model assessment for insulin resistance; MBE=mind-body exercise; RE=resistance exercise; OGTT=oral glucose tolerance test.

**Figure 3.**
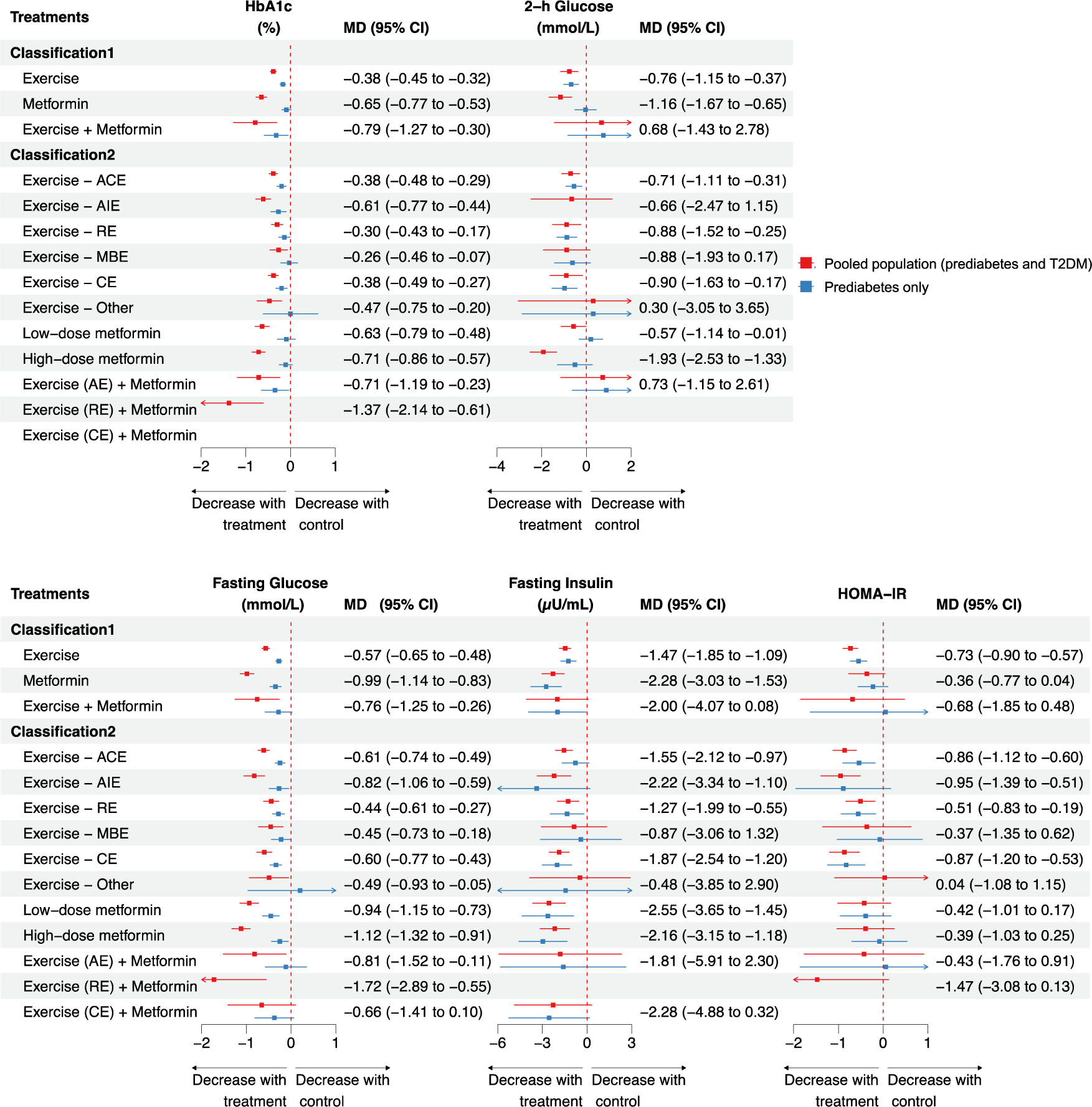
Forest plots of network meta-analysis (indirect comparison) for glucose parameters in pooled population and prediabetes only. *Note.* The MD (95% CI) values in the figure are presented for the pooled population (i.e., both T2DM and prediabetes; in red), but not for the group of “prediabetes individuals only” (in blue; see detailed MD [95% CI] Appendix 12). ACE=aerobic continuous exercise; AIE=aerobic interval exercise; CE=combined exercise; CI=confidence interval; HOMA-IR = homeostatic model assessment for insulin resistance; MD=mean difference; MBE=mind-body exercise; RE=resistance exercise.

**Figure 4.**
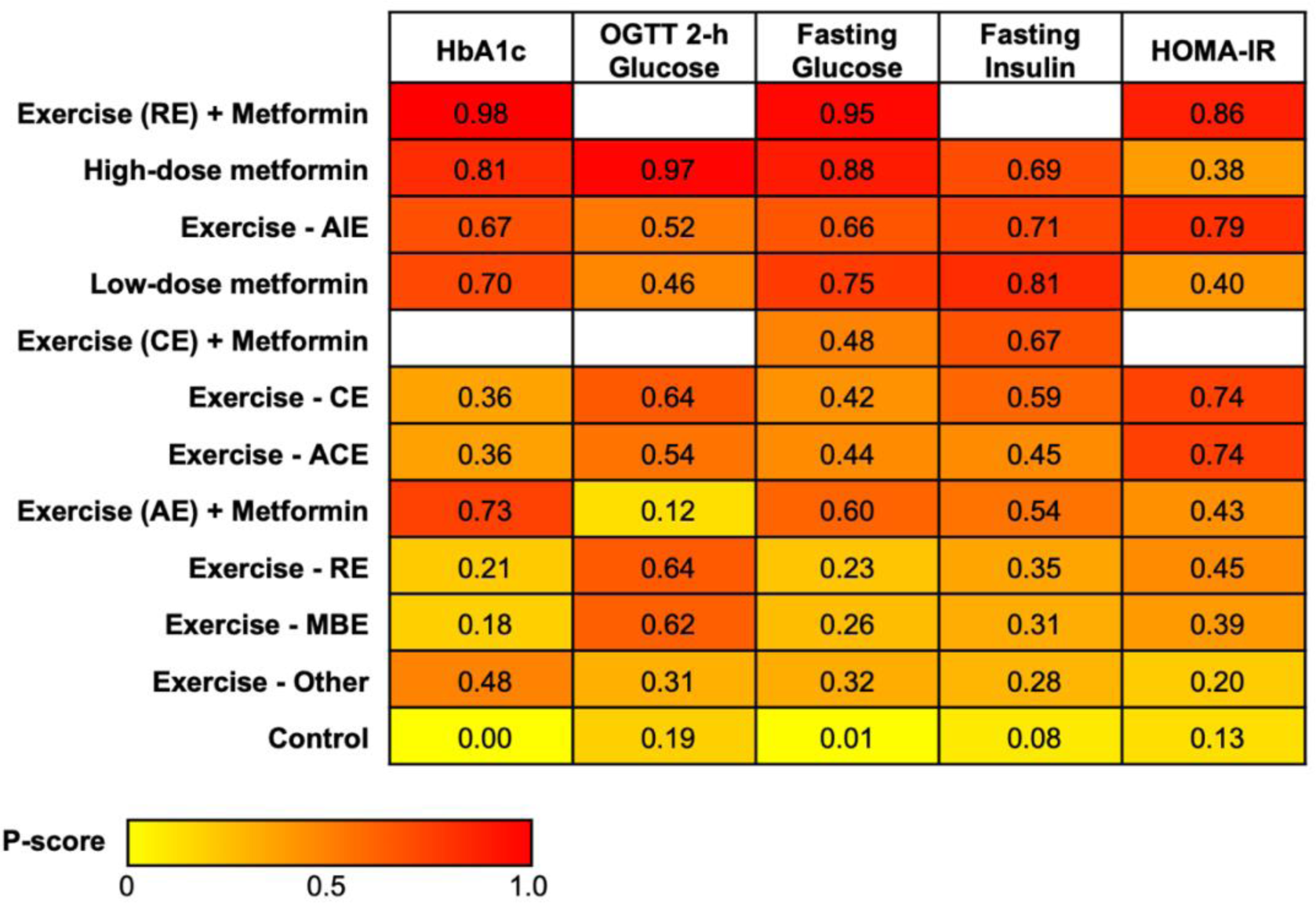
Heat map of treatments ranked according to the associated degree of alteration in glucose profiles. Note. Numbers reflect P-score, which ranks treatments on a continuous scale from 0 to 1. A higher P-score indicates a greater decrease in the glucose metabolic parameter. White squares indicate that data were not available. The treatments are arranged in descending order of their average efficacy on all glucose metabolic parameters, from highest to lowest probability of being the best. ACE=aerobic continuous exercise; AIE=aerobic interval exercise; CE=combined exercise; HOMA-IR=homeostatic model assessment for insulin resistance; MBE=mind-body exercise; RE=resistance exercise; OGTT=oral glucose tolerance test.

### OGTT 2-h glucose

For the changes in 2-h glucose during OGTT of the pooled population, 50 RCTs were included in the network with 33 comparisons for exercise, 17 for metformin, and 1 for exercise + metformin, respectively (Figure 2B). The results of both pairwise and network meta-analysis showed that standalone metformin (Figure 2B, Figure 3, and Appendix 10; −1.16 [−1.67 to −0.65] mmol/L) had larger effects on 2-h glucose than exercise (−0.76 [−1.15 to −0.37] mmol/L). There was no study directly comparing exercise + metformin and control, while network meta-analysis found no evidence of a decrease in 2-h glucose observed with this group (0.68 [−1.43 to 2.78] mmol/L). The network meta-analysis in detailed classification found that aerobic continuous exercise (−0.71 [−1.11 to −0.31] mmol/L), resistance exercise (−0.88 [−1.52 to −0.25] mmol/L), combined exercise (−0.90 [−1.63 to −0.17] mmol/L) as well as low-dose (−0.57 [−1.14 to −0.01] mmol/L) and high-dose metformin (−1.93 [−2.53 to −1.33] mmol/L) decreased 2-h glucose significantly (Figure 3). The ranking on 2-h glucose levels indicated high-dose metformin as the best and aerobic exercise + metformin as the worst treatment (Figure 4; Appendix 13).

### Fasting glucose

For the changes in fasting glucose of the pooled population, 320 RCTs were included in the network with 254 comparisons for exercise, 74 for metformin, and seven for exercise + metformin, respectively (Figure 2C). The results of the pairwise meta-analysis comparing with the control group showed that metformin (Figure 2C and Appendix 10; −1.01 [−1.16 to −0.85] mmol/L) had a larger effect on fasting glucose than exercise (−0.56 [−0.64 to −0.47] mmol/L) and exercise + metformin (−0.30 [−1.37 to 0.77] mmol/L). Furthermore, the network meta-analysis found exercise + metformin (Figure 3 and Appendix 10; −0.76 [−1.25 to −0.26] mmol/L) was more efficacious than exercise alone (−0.57 [−0.65 to −0.48] mmol/L) but less than metformin alone (−0.99 [−1.14 to −0.83] mmol/L). The network meta-analysis found that all modalities of exercise, all dosages of metformin, aerobic exercise + metformin, and resistance exercise + metformin significantly decreased fasting glucose with MD ranging from −1.72 (−2.89 to −0.55) to −0.44 (−0.61 to −0.27) mmol/L (Figure 3). Ranking on fasting glucose level indicated resistance exercise + metformin as the best and resistance exercise alone as the worst (Figure 4; Appendix 13).

### Fasting insulin

For the changes in fasting insulin of the pooled population, 164 RCTs were included in the network with 127 comparisons for exercise, 42 for metformin, and 6 for exercise + metformin, respectively (Figure 2D). The results of the pairwise meta-analysis comparing with the control group showed that exercise + metformin (Figure 2D and Appendix 10; −4.40 [−7.99 to −0.81] µU/mL) had a larger effect on fasting glucose than metformin (−2.33 [−3.09 to −1.56] µU/mL) and exercise alone (−1.49 [−1.87 to −1.11] µU/mL). However, the network meta-analysis showed that exercise + metformin had no significant effect on fasting insulin (Figure 3; −2.00 [−4.07 to 0.08] µU/mL), which was inconsistent with the pairwise meta-analysis. The network meta-analysis in detailed classification found that only continuous aerobic exercise (Figure 3; −1.55 [−2.12 to −0.97] µU/mL), aerobic interval exercise (−2.22 [−3.34 to −1.10] µU/mL), resistance exercise (−1.27 [−1.99 to −0.55] µU/mL), combined exercise (−1.87 [−2.54 to −1.20] µU/mL), low-dose (−2.55 [−3.65 to −1.45] µU/mL) and high-dose metformin (−2.16 [−3.15 to −1.18] µU/mL) significantly decreased fasting insulin. The ranking on the fasting insulin level indicated low-dose metformin as the best and other exercises as the worst treatment (Figure 4; Appendix 13).

#### HOMA-IR

For the changes in HOMA-IR of the pooled population, 118 RCTs were included in the network with 100 comparisons for exercise, 18 for metformin, and 2 for exercise + metformin, respectively (Figure 2E). The results of the pairwise meta-analysis comparing with the control group revealed that the HOMA-IR index decreased significantly with exercise only (Figure 2E and Appendix 10; −0.73 [−0.89 to −0.56]) but not metformin (−0.39 [−0.80 to 0.02]). However, there was no study directly comparing exercise + metformin and control, while the network meta-analysis found no evidence of a decrease in HOMA-IR with exercise + metformin group (Figure 3; −0.68 [−1.85 to 0.48]). In the network meta-analysis for the detailed classification of exercise, only continuous aerobic exercise (Figure 3; −0.86 [−1.12 to −0.60]), continuous interval exercise (−0.95 [−1.39 to −0.51]), resistance exercise (−0.51 [−0.83 to −0.19]), and combined exercise (−0.87 [−1.20 to −0.53]) significantly reduced HOMA-IR. Ranking on the HOMA-IR level indicated resistance exercise + metformin as the best, and other exercises as the worst treatment (Figure 4; Appendix 13).

### Subgroup and sensitivity analyses

We performed subgroup analyses of prediabetes individuals (Figure 3, and Appendix 12), to assess whether exercise is as efficient as metformin in improving glucose metabolism for prediabetes. We found that exercise was overall more efficient than metformin in improving HbA1c (−0.17 [−0.23 to −0.11] and −0.09 [−0.20 to 0.01] %, respectively), 2-h glucose in OGTT (−0.68 [−1.01 to −0.35] and −0.04 [−0.51 to 0.43] mmol/L, respectively), and HOMA-IR (−0.73 [−0.90 to −0.57] and −0.36 [−0.77 to 0.04], respectively). Meanwhile, the effects on fasting glucose were fairly similar between exercise and metformin treatment (−0.27 [−0.34 to −0.20] and −0.35 [−0.47 to −0.22] mmol/L, respectively). Specifically, according to the ranking for treatments in the detailed classification (Appendix 13), combined exercise was shown as the most effective treatment for 2- h glucose (−0.98 [−1.54 to −0.42] mmol/L; ranking 1) and HOMA-IR (−0.83 [−1.24 to −0.41]; ranking 1) and the second most effective for fasting glucose (−0.34 [−0.47 to −0.21]; ranking 2). The results of subgroup analyses for T2DM patients showed a similar pattern to that observed in the pooled populations (see Appendix 11 and 12).

In accordance with the review protocol, we further conducted sensitivity analyses by excluding trials with overall high risk of bias and trials with intervention duration longer than 12 weeks. The results indicated no substantial change (Appendix 18).

## DISCUSSION

This systematic review and network meta-analysis included 378 RCTs with 30,884 individuals who were randomly assigned into exercise, metformin, exercise + metformin, and control groups, while taking into account the exercise modalities and metformin dosage. To our knowledge, this is the first network meta-analysis to compare the efficacy of exercise, metformin, and their combination on glucose metabolism in individuals with impaired glycemic control (i.e., prediabetes and T2DM). In this meta-analysis, we provide a very comprehensive data synthesis of exercise interventions on glucose metabolism, allowing us to compare the pooled effect of different exercise modalities.

Our main findings indicate that metformin was overall more efficient than exercise treatment in improving glycemic control, when pooling all individuals. Especially in patients diagnosed with T2DM, our subgroup analysis found that metformin decreased HbA1c to a larger extent than exercise (MDs −0.90 [−1.07 to −0.72] vs −0.46 [−0.54 to −0.37] %; Appendix 12). Due to the lack of direct comparison between exercise and metformin in previous reviews, data can only be compared to reviews looking at either metformin or exercise alone.^35,36^ Our results of metformin are somewhat in line with a previous work (i.e., MD −0.92 [−1.07 to −0.77] %),^35^ but small inconsistencies do exist related to the magnitude of the MD of exercise in other work (i.e. MDs -0.67 [−0.84 to −0.49] and −0.43 [−0.59 to −0.28] % for supervised and unsupervised exercise, respectively).^36^ The latter inconsistency may possibly be caused by us including both supervised and unsupervised exercise, which is different from previous work.^36^ However, the effect of exercise on glycemic control in our subgroup analysis was almost equal to and even superior to metformin treatment in prediabetes individuals. This finding is in line with a landmark study– Diabetes Prevention Program (DPP), which showed that lifestyle intervention (including exercise training) reduced the incidence of T2DM by 58% but metformin by only 31%.^37^ Therefore, our data further supported Davidson’s viewpoint, that individuals with high risk of diabetes should firstly receive appropriate lifestyle intervention rather than metformin treatment.^13^

HOMA-IR, as the approximate estimate of insulin resistance, was the only outcome that showed a greater benefit from exercise compared to metformin in both individuals with prediabetes and T2DM. This finding could be explained by the different mechanisms of glycemic control from each treatment. Although metformin may enhance insulin sensitivity by affecting fat metabolism, it primarily lowers glucose by reducing hepatic glucose production.^38^ However, exercise directly benefits insulin resistance in the intramuscular glucose transport system, which appears to be a key benefit of exercise interventions for glycemic control.^39^ Despite the lack of a gold standard measurement of insulin resistance, our finding in HOMA-IR confirmed the distinctive efficacy of exercise in reducing insulin resistance.

Our findings also provide a comprehensive comparison regarding the efficacy of different exercise modalities, based on the analyses of 300 included studies involving exercise interventions. Our classification of exercises was based on the predominant effects and goals exhibited by individual exercise modalities, thereby considering mind-body exercise as a separate modality. We found that combined exercise (i.e., exercise training comprising several types of exercise, mainly aerobic + resistance) and aerobic interval exercise might be the most effective exercise modalities for prediabetes and T2DM, respectively. Several meta-analyses have been conducted to investigate the best exercise modality,^18–20^ while the classification of exercise modality in those studies differed from ours, leading to the inconsistency of the results. For example, one recent network meta-analysis categorized exercise into eight modalities (i.e., supervised aerobic, unsupervised aerobic, anaerobic, balance, supervised resistance, unsupervised resistance, combined aerobic and resistance, and flexibility exercise). They found that flexibility exercise was the most effective exercise for reducing fasting glucose (–1.48 [–2.17 to –0.78] mmol/L) and HbA1c (–0.71 [–1.08 to –0.34] %) in T2DM patients, while only 2 studies were included to support this finding.^20^ However, our results on aerobic exercise, resistance exercise, and combined exercise were in line with existing meta-analyses indicating that combined exercise was superior than performing each mode alone.^18,19^ Our findings regarding aerobic interval vs continuous exercise on HbA1c were also consistent with a previous pairwise meta-analysis.^40^ In this study, high-intensity interval training provided a greater benefit to HbA1c than moderate-intensity continuous training (–0.17 [–0.36 to 0.02] %) in prediabetes and T2DM.

The interactive effect between exercise training and metformin is a debatable topic and was highlighted in a recent consensus statement by the American College of Sports Medicine (ACSM).^10^ We found the overall efficacy of combined exercise and metformin treatment was superior to exercise treatment alone but similar or even inferior to metformin alone. Especially, aerobic exercise might blunt the glycemic control effect of metformin. A possible nonadditive effect might be due to aerobic exercise and metformin inducing opposite effects on oxidative capacity. For example, aerobic exercise enhances muscle mitochondrial function,^41^ and it is discussed that metformin may inhibit the complex 1 of the electron transport chain.^42,43^ It is worth noticing that our results showed the efficacy of resistance exercise + metformin was superior to resistance exercise and metformin treatment alone. The possible reason might be that resistance exercise improves glycemic control through mass effect (i.e., increased fat-free mass), which is different from the effects induced by aerobic exercise.^44^ Importantly, our findings do not imply that patients taking metformin are not suggested to perform exercise, considering that exercise is also beneficial to endothelial function, physical fitness, and mental health.^10^ However, because the inverse variance model was used to summarize pooled effects, our results for combined exercise and metformin intervention might be affected by the small sample size, which may reduce the statistical power.^45^ Therefore, future clinical trials are needed and encouraged to further explore how to optimize the interactive effect of exercise and metformin, e.g. by considering the time interval between exercise and metformin administration, exercise program variables (e.g., modality, intensity, frequency), and drug delivery method (e.g., immediate or extended-release).

## Limitations

Our study had some limitations. First, the confidence of evidence was overall low or very low. The main reason for the downgrading of confidence was the heterogeneity between included studies. Although we performed subgroup analysis considering different populations (i.e., prediabetes and T2DM), the heterogeneity showed no important changes. Second, despite the large number of overall included trials, only five involved exercise + metformin interventions. Thus, it was impossible to further classify these studies based on the metformin dose. Hence, the results should be interpreted with caution.

## CONCLUSIONS

To conclude, this network meta-analysis showed that exercise, metformin, and their combination are effective in improving glycemic control in individuals with impaired glycemic control, including prediabetes and T2DM. Metformin seems more effective than exercise in improving glycemic control while exercise plays a unique role in reducing insulin resistance in the pooled population, but their efficacy was comparable in individuals with prediabetes. Concerning the exercise modalities, aerobic interval exercise and combined exercise appear to be most effective for impaired glucose metabolism in the pooled population. Combined exercise and metformin therapy showed non-inferior effects compared to exercise or metformin alone in the pooled population, depending on the specific exercise modalities. Our findings are supposed to be understood as foundation to further enhance clinical guidelines of both pharmaceutical and non-pharmaceutical first-line treatments of prediabetes and T2DM.

## Data Availability

The full dataset is available on request by contacting the corresponding author.

## Acknowledgments

This study was funded by Chinese Scholarship Council (CSC, no. 202206230079). The views expressed are those of the authors and not necessarily those of the CSC.

## Contributors

TZ, CB, SC, WB, and MS conceived and designed the study. TZ, QY, and MS selected the articles and extracted the data, assessed the risk of bias. TZ and JF analyzed the data. TZ, QY, JF, BY accessed and verified the data. TZ wrote the first draft of the manuscript. TZ, JF, CB, SC, WB, and MS interpreted the data and contributed to the writing and revising of the final version of the manuscript. All authors agreed with the results and conclusions of this article.

## Declaration of interests

We declare no competing interests.

## Data sharing

The full dataset is available on request by contacting the corresponding author.

## Notes

### Competing Interest Statement

The authors have declared no competing interest.

